# Inactivated Zoonotic Influenza A(H5N8) Vaccine Induces Robust Antibody Responses Against Recent Highly Pathogenic Avian Influenza Clade 2.3.4.4b A(H5N1) Viruses

**DOI:** 10.1101/2025.02.12.25322044

**Authors:** Oona Liedes, Nina Ekström, Anu Haveri, Anna Solastie, Saimi Vara, Willemijn F. Rijnink, Theo M. Bestebroer, Mathilde Richard, Rory D. de Vries, Erika Lindh, Niina Ikonen, Terhi Laaksonen, Riikka Holopainen, Laura Kakkola, Maija Lappalainen, Ritva K. Syrjänen, Ilkka Julkunen, Hanna Nohynek, Merit Melin

## Abstract

In 2023, Finland faced an outbreak of highly pathogenic avian influenza caused by clade 2.3.4.4b A(H5N1) viruses, which spread from wild birds to fur farms. Vaccinations of individuals at-risk, such as fur and poultry farm workers, veterinarians, and laboratory workers, began in June 2024 using the MF59-adjuvanted inactivated (H5N8) vaccine manufactured by Seqirus (based on clade 2.3.4.4b A/Astrakhan/3212/2020). We investigated antibody responses following a two-dose vaccination regimen in 39 subjects. Vaccination induced comparable levels of functional antibodies both against the vaccine virus and two clade 2.3.4.4b viruses, either associated with outbreaks in fur animals in Finland or cattle in the United States. Upon two doses of the vaccine for previously unvaccinated people, the seroprotection rate against the vaccine virus was 83 % (95 % CI 70-97 %, titer ≥20) and 97 % (95 % CI 90-100 %, titer ≥40) using microneutralization or hemagglutinin inhibition assays, respectively. In a subset of previously H5-vaccinated individuals, the first dose already led to seroprotective titers, indicative of immunological recall. These data show that the vaccine is expected to confer cross-protection against currently circulating H5 clade 2.3.4.4b viruses.

## Introduction

Highly pathogenic avian influenza (HPAI) clade 2.3.4.4b A(H5Nx) viruses have been expanding their geographic and host range since 2020. These viruses currently cause outbreaks in wild birds and poultry in Asia, Europe, Africa, the Americas and Antarctica, and spillover to mammals at an alarming frequency. Extensive circulation in multiple species already resulted in the acquisition of several traits associated with increased zoonotic potential^1^.

In 2023, Finland experienced a widespread outbreak of clade 2.3.4.4b A(H5N1) that caused mass mortalities among wild birds but also spread to several dozen fur animal farms^2^. The outbreak was seeded by multiple introductions from surrounding wild birds and resulted in the culling of approximately half a million fur animals over a period of 6 months, mainly foxes (*Vulpes vulpes*), arctic foxes (*Vulpes lagopus*), and minks (*Neovison vison*). The causative virus was associated with significant mortality among wild and captive birds across Europe in the same year^2–4^. Epidemiological and genomic investigations indicated several modes of transmission, including environmental contamination, mammal-to-mammal and possibly mechanical transmission. The multiplicity of potent transmission modes posed considerable challenges for mitigation and control efforts through biosecurity measures^3^. Molecular analyses of the A(H5N1) viruses isolated from fur animals revealed multiple amino acid changes in PB2 and neuraminidase (NA) proteins, which have been associated with adaptation to mammalian hosts^2,3^. Despite numerous exposure events among different occupational groups involved in outbreak control, no human infections were detected.

The expanding host range of clade 2.3.4.4b A(H5N1) viruses was further exemplified in March 2024, when an outbreak was reported in dairy cattle in the United States. The outbreak was seeded through a single introduction from wild birds and is believed to be spread and maintained mainly through mechanical transmission (e. g. animal movements and contaminated milking equipment)^5^. After more than one year of sustained multi-state circulation, the outbreak is still not under control and has been associated with numerous human cases (https://www.cdc.gov/bird-flu/situation-summary/index.html).

In 2024, a total of 72 human cases were reported globally, which included cases from the United States as well as from Australia, Cambodia, Canada, China, and Vietnam^6^. Most cases were linked to exposure to sick or dead animals, or their excretions. Disease severity in humans varied from mild to fatal. High fatality rates were previously reported for A(H5N1) cases, but in 2024, the case fatality rate was around 5 % with most cases presenting with mild disease. Mild cases were mostly identified through surveillance, suggesting that zoonotic transmission could be more common than previously thought^1,6^. Several serological studies also suggest that there is a low level of zoonotic transmission of H5N1 among exposed occupational groups^1^. Considering the number of reared animal populations that have been affected by H5N1 and the number of people who are consequently exposed, human cases remain rare.

The global distribution, establishment of endemic circulation in wild birds, poultry and mammals, and expanding host range of HPAI clade 2.3.4.4b A(H5Nx) viruses raise concerns over future zoonotic events and establishment of circulation in humans. Infection of humans could lead to mutations in the virus receptor-binding preferences, as observed in recent cases reported from Canada and the US (https://www.cdc.gov/bird-flu/spotlights/h5n1-response-12232024.html)^7^. Additionally, human infection leads to increased opportunity for reassortment with other influenza A viruses. Together, this demonstrates the risk of acquiring pandemic traits through human adaptive evolution.

Implementation of biosecurity measures and safe farming practices to mitigate risk to humans was met with substantial resistance during the fur farm outbreak in Finland. Due to the continued practice of fur farming in Finland, and the need to increase occupational safety in the context of the developing epizootic situation, it was decided to offer an A(H5N8) influenza vaccine to individuals considered at risk of exposure to HPAI (workers at fur farms, poultry workers, public sector veterinarians, bird ringers, and laboratory personnel handling A(H5) HPAI viruses or samples that may contain these viruses)^8,9^. Finland acquired the MF59-adjuvanted zoonotic influenza vaccine based on A/Astrakhan/3212/2020 (H5N8, clade 2.3.4.4b), manufactured by Seqirus, as part of the European Union’s joint procurement agreement. This vaccine is expected to provide cross-protection against currently circulating clade 2.3.4.4b viruses (https://www.ema.europa.eu/en/medicines/human/EPAR/zoonotic-influenza-vaccine-seqirus). Vaccination efforts in Finland commenced on June 13, 2024, as soon as the vaccine received marketing authorisation from the European Medical Agency (EMA) (https://ec.europa.eu/health/documents/community-register/html/h1761.htm).

To date, Finland remains the only country that started vaccinating risk groups with this vaccine. There are no data of vaccine immunogenicity or immune protection of this vaccine against clade 2.3.4.4b influenza viruses in humans. Since HPAI clade 2.3.4.4bA(H5N1) viruses show genetic variability in the hemagglutinin (HA) protein, which could lead to antigenic changes influencing vaccine effectiveness, it is essential to study protection against viruses from recent and current outbreaks alongside the vaccine strain. In this study, we investigated vaccine-induced antibody responses in at-risk individuals who were offered the vaccine in Finland in 2024. We measured functional and binding antibodies targeting both the vaccine virus and circulating clade 2.3.4.4b A(H5N1) viruses with microneutralization (MN) and hemagglutinin inhibition (HI) assays and with fluorescent bead-based multiplex immunoassay (FMIA). Some of the participants had previously received A(H5N1) vaccines offered to a limited target group in 2009, 2011-2012 and/or 2018, allowing for the evaluation of immunological recall responses.

## Results

### Study population

We enrolled 52 participants in the study in July-September 2024 (**Figure 1A**). Most of the study participants were laboratory personnel (n=31) and the second largest group consisted of bird ringers (n=12). Only a small number of poultry workers (n=5) and veterinarians (n=4) participated in the study. Although individuals working on fur farms were the primary target group for the vaccine, none participated. Blood samples were obtained before vaccination and after each of the vaccine doses (**Figure 1B**). Of the 52 participants, 40 provided serum samples at all three study visits. The age of the participants ranged from 27 to 77 years, and 73 % were female. Since vaccine-induced immune responses are affected by age, older adults typically exhibiting a reduced immune response, we limited our analysis to the age group ≤65 years and to participants who provided blood samples at scheduled visits. In total, samples from 39 participants were included (**Figure 1A**, **Table 1**). The vaccine doses were administered at a median interval of 28 days. The baseline serum sample was obtained at a median of 3 days before vaccination and the post-vaccination samples at a median of 20 and 21 days after the first and second vaccine dose, respectively (**Figure 1B**). Nine study participants had previously received two to six doses of A(H5N1) vaccines in 2009, 2011-2012 and/or 2018 (**Tables 1 and 2**).

**Figure 1.**
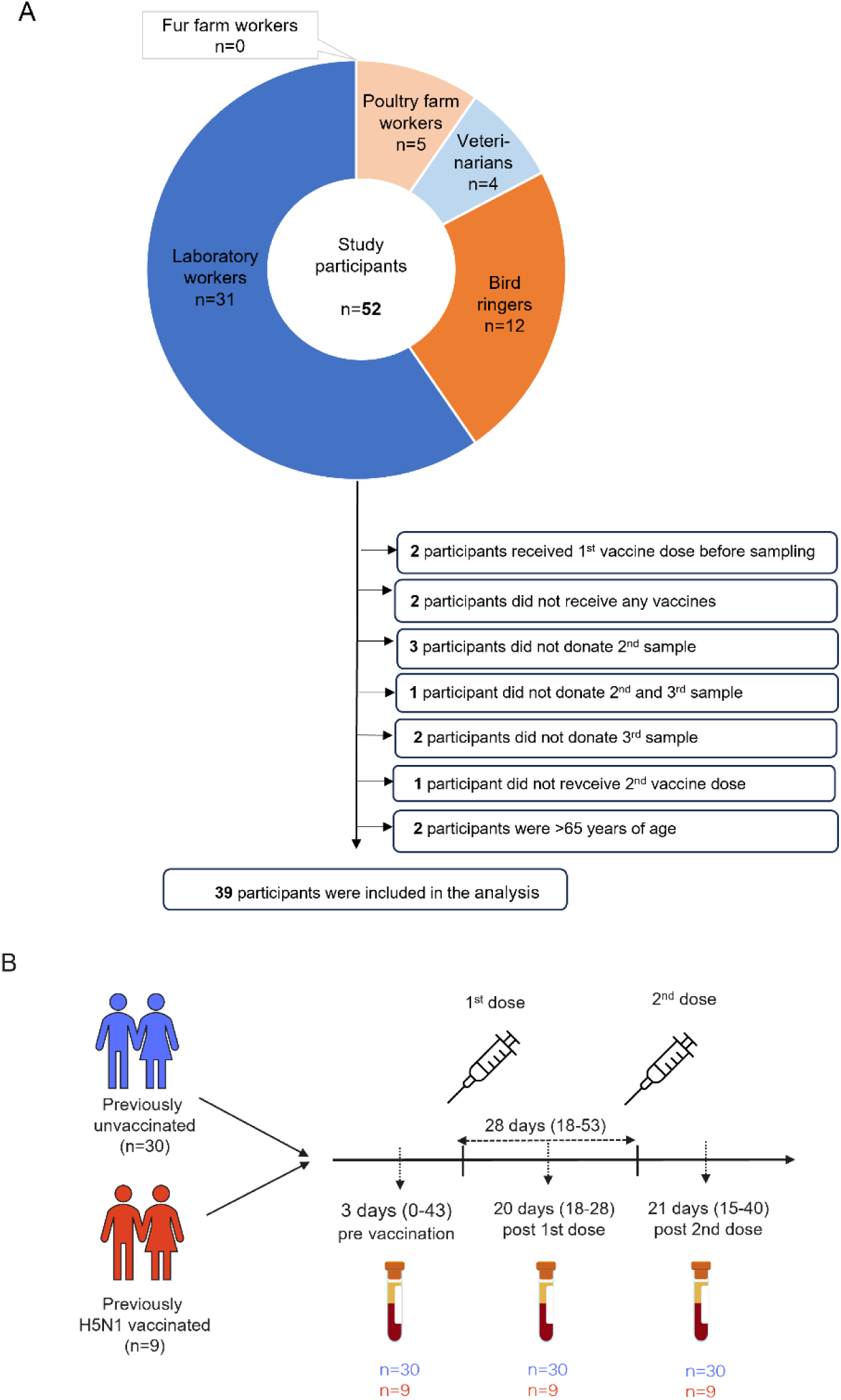
Study overview. (A) Flow chart showing the study population by vaccine target groups and reasons for exclusion from analysis. (B) Timeline of vaccinations and samplings in the two study groups of previously unvaccinated (n=30) and A(H5N1) vaccinated (n=9) participants. The MF59-adjuvanted A(H5N8) influenza vaccine (clade 2.3.4.4b A/Astrakhan/3212/2020, Seqirus) was administered as a two-dose regimen with a median of 28 days (range for dose interval is indicated between brackets). Blood was collected before vaccination, after the first and second vaccine dose (median and range of days shown).

**Table 1.** Characteristics of study participants.

|  | **Previously**<br>**unvaccinated** | **Previously**<br>**vaccinated** |
| --- | --- | --- |
| **n** | 30 | 9 |
| **Gender, n (%) female** | 24 (80 %) | 8 (89 %) |
| **Age (years), median (range)** | 41 (27–63) | 51 (40–61) |
| **Target group** |  |  |
| Fur farm workers | 0 (0 %) | 0 (0 %) |
| Poultry farm workers | 3 (10 %) | 0 (0 %) |
| Veterinarians | 4 (13 %) | 0 (0 %) |
| Bird ringers | 5 (17 %) | 0 (0 %) |
| Laboratory workers | 18 (60 %) | 9 (100 %) |
| **Dose interval (days), median (range)** | 28 (20-53) | 28 (18-42) |
| **Number of previous H5 vaccine doses, n (%)** |  |  |
| 2 | NA | 4 (45 %) |
| 4 | NA | 2 (22 %) |
| 6 | NA | 3 (33 %) |

NA= not applicable

### Antibody responses targeting the vaccine antigen

We measured functional antibodies targeting A/Astrakhan/3212/2020 antigen using the MN and the HI assays. Before vaccination, none of the previously unvaccinated participants had measurable neutralizing antibodies (MN ≥10) (**Figure 2A**, **Table 3**). The seroprotection rates (SPRs) were defined as the percentage of participants with MN titer ≥20 and/or percentage of participants with HI titer ≥40. Six of 30 participants had detectable HI antibodies (HI ≥10), but none attained the level of seroprotection^10^ (**Figure 2B**, **Table 3**). In contrast, of the participants who had been previously vaccinated with an H5 vaccine (**Table 1**, **Table 2**), two out of nine had detectable neutralizing antibodies, and seven out of nine had detectable HI antibodies. Notably, one participant reached seroprotection based on HI titers.

**Figure 2.**
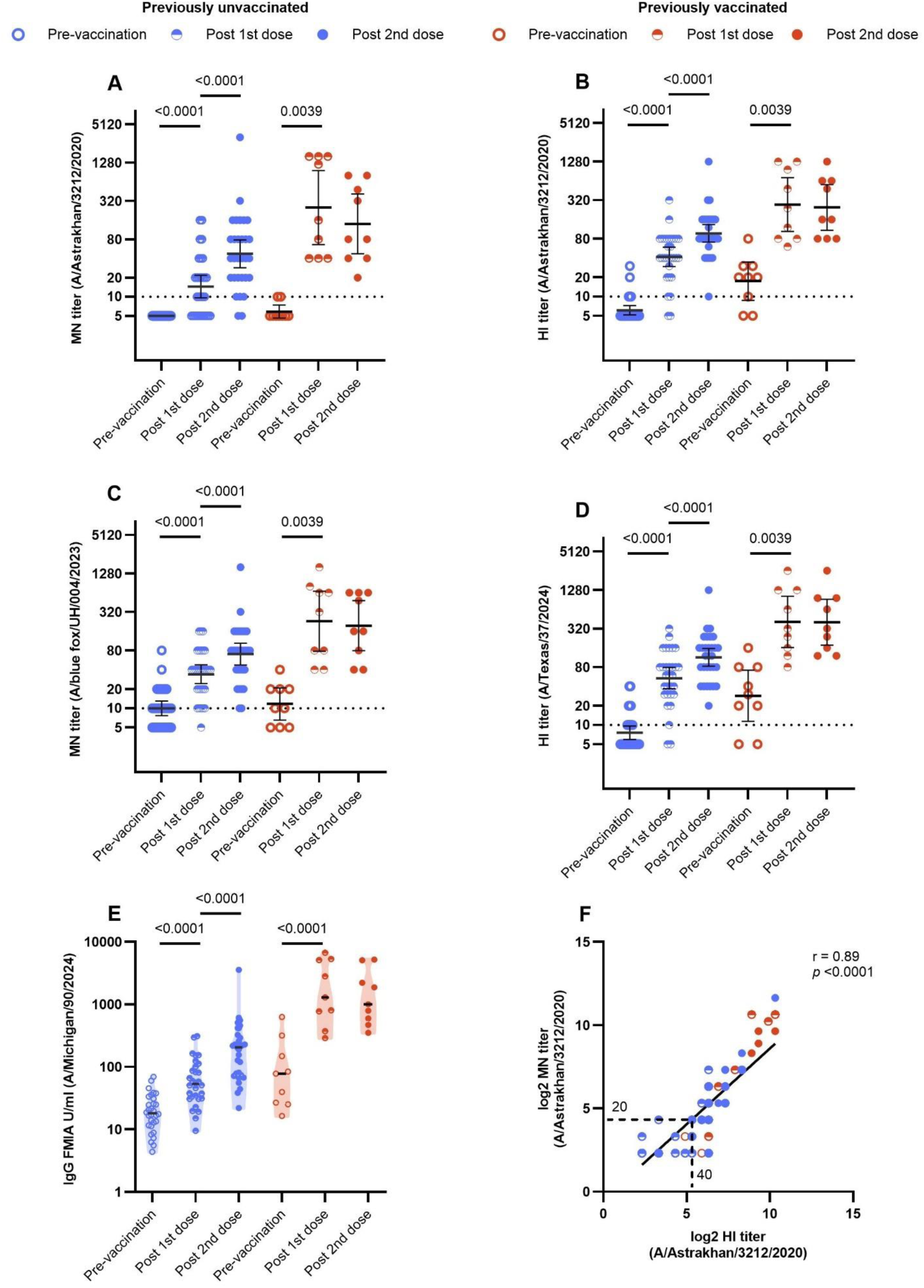
Antibody responses targeting the vaccine antigen and heterologous A(H5N1) viruses. (A, B) Antibodies targeting the vaccine antigen A(H5N8) A/Astrakhan/3212/2020 were measured using the microneutralization (MN) assay and the hemagglutination inhibition (HI) assay. (C) Antibodies targeting A(H5N1) A/blue fox/UH/004/2023 were measured by MN assay. (D) Antibodies targeting A(H5N1) A/Texas/37/2024 antigen were measured by HI assay. (E) HA-specific IgG antibodies binding A(H5N1) A/Michigan/90/2024 were measured by fluorescent bead-based multiplex immunoassay (FMIA). Data was categorized into two groups: A(H5N1) unvaccinated (n=30) and previously vaccinated (n=9), at three time points: pre-vaccination, and three weeks after the first dose and the second dose. The graphs display geometric mean titers (GMT) and concentrations (GMC) (lines) and 95 % confidence intervals (whiskers). The dashed line indicates the positivity threshold, a titer of 10 or above considered positive. Only statistically significant differences between time points within groups are indicated. Comparisons within a group between two time points were conducted using two-tailed Wilcoxon matched-pairs signed-rank test for MN and HI data, and the T-test for FMIA data. (F) The correlation between MN and HI antibody titers to the vaccine antigen A/Astrakhan/3212/2020 was assessed by simple linear regression on log2-transformed data. Each dot (n=142) may represent values from multiple participants.

**Table 2.** A(H5) vaccines administered in previously vaccinated participants.

|  | Pre-pandemic<br>A(H5N1),<br>inactivated,<br>AS03-adjuvanted<br>A/Indonesia/5/200<br>5-like split virion<br>vaccine,<br>clade 2.1.3.2<br>(GlaxoSmithKline) |  | A(H5N1)<br>inactivated<br>adjuvant-free<br>A/Vietnam/1203/2<br>004-like whole<br>virus vaccine,<br>clade 1 (Baxter) |  | A(H5N1) inactivated<br>MF59-adjuvanted<br>A/turkey/Turkey/1/2<br>005 (H5N1)-like<br>strain (NIBRG-23)<br>vaccine,<br>clade 2.2.1<br>(Novartis) |  | Zoonotic A(H5N8),<br>inactivated, M59-adjuvanted,<br>A/Astrakhan/3212/2<br>020-like strain (CBER-RG8A) vaccine,<br>(clade 2.3.4.4b)<br>(Seqirus Vaccines) |  |
| --- | --- | --- | --- | --- | --- | --- | --- | --- |
|  | **2009** |  | **2011-2012** |  | **2018** |  | **2024** |  |
| **Participant** | 1<sup>st</sup> dose | 2<sup>nd</sup> dose | 1<sup>st</sup> dose | 2<sup>nd</sup> dose | 1<sup>st</sup> dose | 2<sup>nd</sup> dose | 1<sup>st</sup> dose | 2<sup>nd</sup> dose |
| 1 |  |  |  |  | • | • | • | • |
| 2 |  |  |  |  | • | • | • | • |
| 3 | • | • | • | • |  |  | • | • |
| 4 | • | • | • | • | • | • | • | • |
| 5 | • | • | • | • | • | • | • | • |
| 6 | • | • | • | • | • | • | • | • |
| 7 |  |  |  |  | • | • | • | • |
| 8 | • | • |  |  | • | • | • | • |
| 9 |  |  |  |  | • | • | • | • |

Detailed information and the number of A(H5) vaccine doses given prior to and during this study indicated by year for each participant. Vaccination is indicated by a solid circle (•).

**Table 3.** Antibody responses to the A(H5N8) vaccine antigen measured by the microneutralization (MN) and the hemagglutination inhibition (HI) assays.

|  |  | Previously unvaccinated (n=30) |  |  | Previously vaccinated (n=9) |  |  |
| --- | --- | --- | --- | --- | --- | --- | --- |
|  |  | Pre-vaccination | 3 weeks post 1<sup>st</sup> dose | 3 weeks post 2<sup>nd</sup> dose | Pre-vaccination | 3 weeks post 1<sup>st</sup> dose | 3 weeks post 2<sup>nd</sup> dose |
| **MN** | **GMT [95 % CI]** | 5.00 [5.00-5.00] | 14.5 [9.64-21.7] | 47.4 [28.7-78.3] | 5.83 [4.61-7.38] | 252 [65.8-962] | 140 [47.4-412] |
|  | **% positive ≥ 1:10 (n/n)** | 0 % (0/30) | 63 % (19/30) | 93 % (28/30) | 22 % (2/9) | 100 % (9/9) | 100 % (9/9) |
|  | **% seropositive ≥ 1:20 (n/n)** | 0 % (0/30) | 47 % (14/30) | 83 % (25/30) | 0 % (0/9) | 100 % (9/9) | 100 % (9/9) |
| **HI** | **GMT [95 % CI]** | 6.10 [5.15-7.21] | 41.7 [29.5-58.9] | 96.6 [70.2-133] | 17.4 [8.74-34.5] | 273 [104-718] | 246 [108-560] |
|  | **% positive ≥ 1:10 (n/n)** | 20 % (6/30) | 93 % (28/30) | 100 % (30/30) | 78 % (7/9) | 100 % (9/9) | 100 % (9/9) |
|  | **% seropositive ≥ 1:40 (n/n)** | 0 % (0/30) | 73 % (22/30) | 97 % (29/30) | 11 % (1/9) | 100 % (9/9) | 100 % (9/9) |

GMT = geometric mean titer, CI = confidence interval

In previously unvaccinated participants, a single vaccine dose induced functional antibodies targeting the A/Astrakhan/3212/2020 antigen, with geometric mean titers (GMTs) of 14.5 or 41.7 measured by MN or HI respectively (**Figure 2A and 2B**, **Table 3**). The antibody levels increased 2.89-fold for MN and 6.83-fold for HI (**Figure 3A and 3B**). After one dose, 47 % (95 % CI: 29-65 %) of the previously unvaccinated participants reached the seroprotection level based on MN, and 73 % (95 % CI 58-89 %) based on HI. Following the second dose, antibody levels increased 3.27-fold or 2.32-fold compared to antibody levels after the first dose as determined by MN or HI, respectively. SPRs after the second dose were 83 % (95 % CI: 70-97 %) and 97 % (95 % CI: 90-100 %) based on MN and HI, respectively.

**Figure 3.**
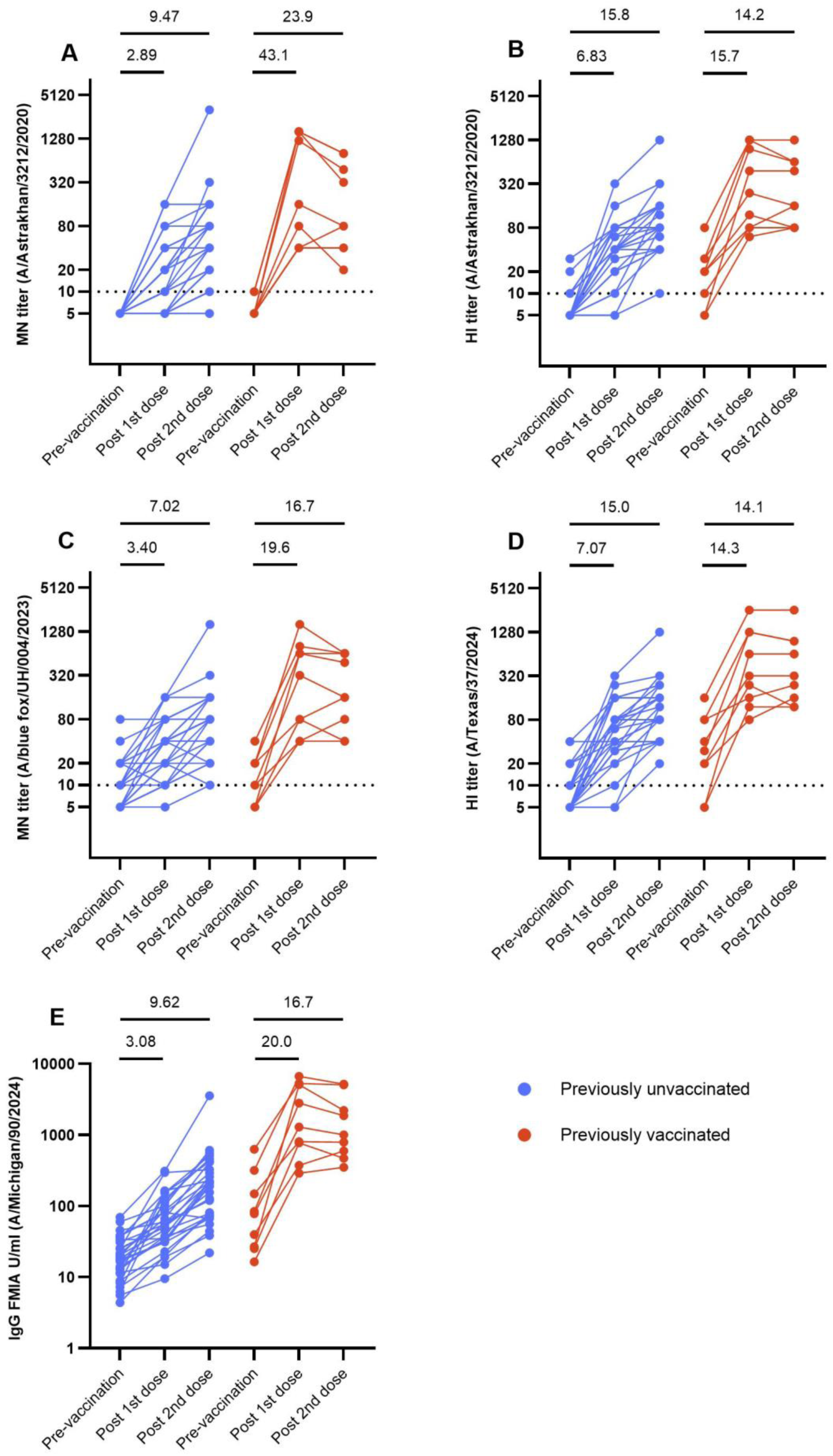
Kinetics of vaccine-induced antibody responses. (A,B) Antibodies targeting the vaccine antigen A(H5N8) A/Astrakhan/3212/2020 were measured using the microneutralization (MN) and the hemagglutination inhibition (HI) assay. (C) Antibodies targeting A(H5N1) A/blue fox/UH/004/2023 were measured by the MN assay. (D) Antibodies targeting A(H5N1) A/Texas/37/2024 were measured by HI assay. (E) HA-specific IgG antibodies binding A(H5N1) A/Michigan/90/2024 were measured by fluorescent bead-based multiplex immunoassay (FMIA). Individual responses are shown as lines for two groups, A(H5N1) unvaccinated (n=30) and previously vaccinated (n=9), at three time points: pre-vaccination, and three weeks after the first dose and the second dose. The dashed line indicates the positivity threshold. Fold changes before vaccination and after the first dose, as well as before vaccination and after the second dose, represented by lines within the graphs.

In participants who had previously received an A(H5N1) vaccine, a single vaccine dose induced functional antibodies targeting A/Astrakhan/3212/2020 antigen, GMTs of 252 or 273 measured by MN or HI, respectively (**Figure 2A and 2B**, **Table 3**). The antibody levels increased 43.1-fold for MN and 15.7-fold for HI (**Figure 3A and 3B**, **Table 3**). Antibody levels were significantly higher compared to levels in previously unvaccinated participants, (*p*<0.0001, Mann–Whitney U test) for both MN and HI. After one dose, 100 % (95 % CI: 66-100 %) of the previously vaccinated participants reached the seroprotection level based on both MN and HI. After the second dose, antibody levels did not significantly differ compared to the first dose (MN: *p*=0.0781, HI: *p*=0.4688, Wilcoxon matched-pairs signed-rank test). SPRs after the second dose remained at 100 % (95 % CI: 66-100 %) based on both MN and HI.

A strong positive correlation between the titers measured by MN and HI was observed (r=0.89, *p*<0.0001, Spearman’s correlation) (**Figure 2F**).

### Antibody responses targeting A(H5N1) viruses from recent outbreaks

To assess immune responses against viruses associated with recent outbreaks in mammals, we measured antibody responses targeting two heterologous A(H5N1) HPAI viruses. Antibody titers against A/blue fox/UH/004/2023 virus, isolated from a fur farm in Finland, were determined by MN, and those against A/Texas/37/2024 virus, isolated from a dairy farmer in the United States, were determined by HI (**Table 4**, **Table 5**, **Figure 2C** and **2D**). In addition to MN and HI, we also measured HA-specific binding IgG against A/Michigan/90/2024 (**Table 4**) using FMIA.

**Table 4.** Viruses and antigens used in the microneutralization (MN) assay, the hemagglutination inhibition (HI) assay, and the fluorescent bead-based multiplex immunoassay (FMIA).

| Virus or antigen name | GISAID isolate ID | HA accession number | Subtype | HA clade | Full virus or RG or HA | Passage history | Origin of the virus or HA | Biosafety level handling | Method |
| --- | --- | --- | --- | --- | --- | --- | --- | --- | --- |
| A/Astrakhan/321 2/2020 CVV | EPI_ISL_13655139 | EPI2084527 | A(H5N8) | 2.3.4.4.b | IDCDC-RG71A | E1M2 | The Crick Worldwide Influenza Centre, London (Ex/E1) | BSL2+ | MN |
| A/Astrakhan/321 2/2020 | EPI_ISL_1038924 | EPI1846961 | A(H5N8) | 2.3.4.4.b | 7+1 PR/8 HY | 293TM2 | Gene synthesis from Proteogenix | BSL2 | HI |
| A/blue fox/UH/004/2023 | EPI_ISL_18764855 (E1 passage) | EPI2914899 (E1 passage) | A(H5N1) | 2.3.4.4.b | Full virus | M2 | Associate Professor Tarja Sironen, University of Helsinki | BSL3 | MN |
| A/Texas/37/2024 | EPI_ISL_19027114 | EPI3171488 | A(H5N1) | 2.3.4.4.b | 7+1 PR/8 HY | 293TM2 | Plasmid from Dr. Daniel R. Perez, University of Georgia, USA | BSL2 | HI |
| A/Michigan/90/2 024 | EPI_ISL_19162802 | EPI3334182 | A(H5N1) | 2.3.4.4.b | HA amino acids 1-530 | Original | Antigen from Native Antigen Company | BSL1 | FMIA |

CVV = candidate vaccine virus, HA = hemagglutinin, RG = reverse genetics, PR/8 HY = A/Puerto Rico/8/1934 high yield, M = MDCK.

**Table 5.**
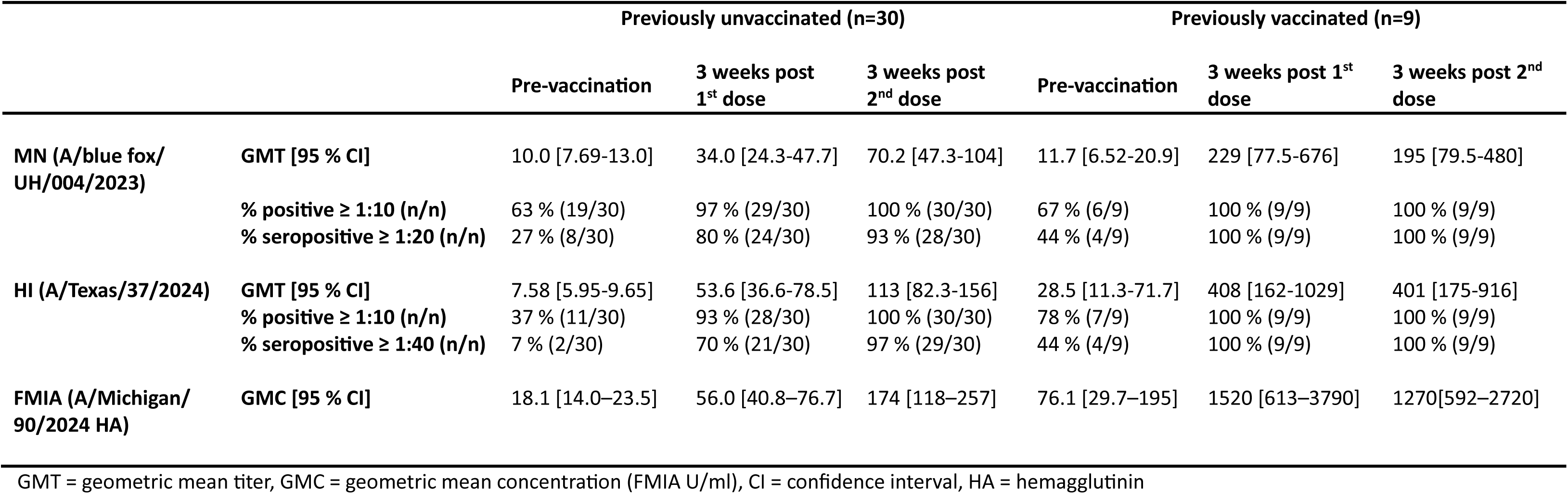
Cross-reactivity of vaccine-induced antibodies with heterologous A(H5N1) viruses measured by the microneutralization (MN) assay, the hemagglutination inhibition (HI) assay, and the fluorescent bead-based multiplex immunoassay (FMIA).

|  |  | Previously unvaccinated (n=30) |  |  | Previously vaccinated (n=9) |  |  |
| --- | --- | --- | --- | --- | --- | --- | --- |
|  |  | Pre-vaccination | 3 weeks post 1<sup>st</sup> dose | 3 weeks post 2<sup>nd</sup> dose | Pre-vaccination | 3 weeks post 1<sup>st</sup> dose | 3 weeks post 2<sup>nd</sup> dose |
| **MN (A/blue fox/**<br>**UH/004/2023)** | **GMT [95 % CI]** | 10.0 [7.69-13.0] | 34.0 [24.3-47.7] | 70.2 [47.3-104] | 11.7 [6.52-20.9] | 229 [77.5-676] | 195 [79.5-480] |
|  | **% positive ≥ 1:10 (n/n)** | 63 % (19/30) | 97 % (29/30) | 100 % (30/30) | 67 % (6/9) | 100 % (9/9) | 100 % (9/9) |
|  | **% seropositive ≥ 1:20 (n/n)** | 27 % (8/30) | 80 % (24/30) | 93 % (28/30) | 44 % (4/9) | 100 % (9/9) | 100 % (9/9) |
| **HI (A/Texas/37/2024)** | **GMT [95 % CI]** | 7.58 [5.95-9.65] | 53.6 [36.6-78.5] | 113 [82.3-156] | 28.5 [11.3-71.7] | 408 [162-1029] | 401 [175-916] |
|  | **% positive ≥ 1:10 (n/n)** | 37 % (11/30) | 93 % (28/30) | 100 % (30/30) | 78 % (7/9) | 100 % (9/9) | 100 % (9/9) |
|  | **% seropositive ≥ 1:40 (n/n)** | 7 % (2/30) | 70 % (21/30) | 97 % (29/30) | 44 % (4/9) | 100 % (9/9) | 100 % (9/9) |
| **FMIA (A/Michigan/**<br>**90/2024 HA)** | **GMC [95 % CI]** | 18.1 [14.0–23.5] | 56.0 [40.8–76.7] | 174 [118–257] | 76.1 [29.7–195] | 1520 [613–3790] | 1270[592–2720] |

GMT = geometric mean titer, GMC = geometric mean concentration (FMIA U/ml), CI = confidence interval, HA = hemagglutinin

Prior to vaccination, 19 out of 30 of previously unvaccinated participants had detectable antibodies targeting A/blue fox/UH/004/2023, and 11 out of 30 had detectable antibodies targeting A/Texas/37/2024. In previously unvaccinated participants, a single vaccination increased antibody levels by 3.40-fold against A/blue fox/UH/004/2023 in MN (GMT 34.0), and by 7.07-fold against A/Texas/37/2024 in HI (GMT 53.6) (**Figure 2C and 2D**, **Table 5**). A single vaccine dose induced geometric mean concentration (GMC) 56 FMIA U/ml of HA-specific IgG antibodies binding A/Michigan/90/2024 (**Figure 2E**). The second vaccine dose increased antibody levels by 2.06-fold with the MN assay (GMT 70.2), 2.11-fold with the HI assay (GMT 113) and 3.08-fold in the FMIA assay (GMC 174 FMIA U/ml) (**Figure 3C-E**).

Of the previously vaccinated participants, six out of nine had measurable MN titers and seven out of nine had measurable HI titers prior to vaccination. Previously A(H5N1)-vaccinated had higher baseline IgG levels (76 FMIA U/ml) compared to the unvaccinated (18 FMIA U/ml, *p*=0.0077, t-test). A single vaccine dose increased antibody levels by 19.6-fold in the MN assay (GMT 229), 14.3-fold in the HI assay (GMT 408) and 20.0-fold in the FMIA assay (GMC 1520 FMIA U/ml) (**Figure 3C-E**). Similar to what was observed with the vaccine antigen, the antibody levels did not significantly differ after the first and the second dose (MN: *p*=0.4531, HI: *p*>0.9999, Wilcoxon matched-pairs signed-rank test).

## Discussion

In this study, we evaluated the immunogenicity of the zoonotic influenza vaccine based on clade 2.3.4.4b A(H5N8) influenza virus manufactured by Seqirus. Finland was the first country to implement vaccination to protect at-risk occupational groups. We had a unique opportunity, and an imperative, to study the immune responses generated by the vaccine in the context of its deployment.

We found that a two-dose series vaccination regimen induced good antibody responses, recognizing both the vaccine virus and other clade 2.3.4.4b A(H5N1) viruses associated with recent outbreaks in fur animals in Finland and cattle farms in the United States. As expected with a primary vaccination, seroprotection of all participants against the vaccine antigen was not observed: 3 % of participants had antibody levels below the seroprotection threshold according to the HI assay, 17 % according to the MN assay.

While the immunogenicity of this vaccine has not been studied in humans before, seroprotection rates against vaccine viruses included in previous vaccine compositions have been assessed in clinical trials^11^. In prior studies with the A(H5N1) A/Vietnam/1194/2004 (clade 1) vaccine and A(H5N1) A/turkey/Turkey/1/2005 (clade 2.2.1) vaccine, seroprotection rates ranged from 85 % (79-91) to 91 % (87-95) for A/Vietnam, and were 91 % (85-94) for A/Turkey. HA-specific antibody levels in the clinical trials were measured by single radial hemolysis (SRH). In the clinical trials MN results against homologous A/Vietnam/1194/2004 indicated seroprotection rates of 67 % (60-74) and 65 % (58-72) in the two studies. In the third study, MN results against homologous A/turkey/Turkey/1/2005 indicated seroprotection rate of 85 % (79-90).

It is important to note that our estimate of seroprotection is subject to uncertainties due to a smaller-than-expected sample size. This affected the width of the confidence intervals (70-97 % with MN and 90-100 % with the HI assay). We had calculated that we would need at least 288 participants to accurately assess the seroprotection rate, but due to the low uptake for the vaccine in Finland, we were only able to recruit 30 previously unvaccinated participants. With the target number of participants, we estimated that our results would be within 5 % of the true seroprotection rate. We plan to continue recruiting for the study in the spring of 2025, to achieve more precise estimates of seroprotection.

The MN, HI and FMIA assays measure different types of antibodies and provide distinct insights into the immune response. While the HI assay measures antibodies that inhibit binding to cell surface receptors, the MN assay measures antibodies that prevent the virus from infecting cells. MN can thus be considered more specific in detecting functional antibodies. IgG antibodies measured with the FMIA detect antibodies that bind to any accessible epitopes in the whole HA antigen and can therefore detect also more cross-reactive antibodies that may have been induced by prior vaccinations or infections with influenza viruses. Neutralizing A(H5N1) antibody responses have usually confirmed the trend seen in the HI assay^12–14^, however, the results do not always correlate^15^. In this study, we found a strong correlation between the MN and HI titers (r=0.89). However, differences in the seroprotection rates calculated based on the two assays were observed. Increased sensitivity and higher HI antibody responses have been observed using horse erythrocyte-based HI assay when compared to using turkey or chicken erythrocytes when performing serology of human sera against avian influenza viruses^16,17^. Accordingly, we found that HI titers against the vaccine antigen were higher than those measured with the MN assay, which contradicts earlier findings that have suggested that the MN assay is a more sensitive method than the HI assay^18^. A theoretical protective value of MN antibody titer for influenza has not been established but it varies depending on the viruses and methods used^19–21^. In addition, virus dilution used and expression of the initial serum dilution vary among laboratories which influence absolute HI and MN titers^18^.

This study included a small number of laboratory workers who had previously received different H5 vaccines in 2009, 2011-2012, and/or 2018 (A/Indonesia/5/2005, clade 2.1.3.2, GlaxoSmithKline, A/Vietnam/1203/2004, clade 1, Baxter, and A/turkey/Turkey/1/2005, clade 2.2.1, Novartis, respectively). In these previously vaccinated participants, a single vaccination elicited a strong antibody response. The second dose did not boost the antibody response further. This finding is consistent with our previous study involving individuals vaccinated in 2009 and 2011-2012^22^. A second dose given three weeks after the first did not significantly increase antibody levels, unlike a subsequent dose given two years later. This suggests that a sufficiently long interval between doses is beneficial for a memory B cell response to mature, and to thereby achieve a proper booster response. This could also contribute to the breadth of the response; when antibody levels are boosted significantly, their combined ability to neutralize viruses different from the vaccine virus is improved. A recent study showed that vaccination with previously licensed, adjuvanted H5N1 vaccines generated neutralizing antibodies that cross-neutralized the A/Astrakhan/3212/2020, clade 2.3.4.4b^23^. Seroprotection rates against Astrakhan after two vaccine doses of AS03-adjuvanted A/Indonesia/05/2005, clade 2.1 were 63.6 % with HI (titer ≥40) and 77.3 % with MN (titer ≥60), and after three doses of MF59-adjuvanted A/Vietnam/1194/2004, clade 1 they were 60 % with HI and 95 % with MN. Findings from another study indicated that vaccination of subjects primed six years prior with the MF59-adjuvanted clade 0-like A/duck/Singapore/1997 (H5N3) vaccine, using a vaccine based on the HA of A/Vietnam/1194/2004, clade 1, induced a higher frequency of circulating memory B cells reactive to A/Vietnam/1194/2004 compared to unprimed subjects^24^. The previously primed subjects had high titers of neutralizing antibodies to antigenically diverse clade 0, 1, and 2 H5N1 viruses as early as day seven. These findings suggest that distant priming can induce a pool of memory B cells that can be rapidly boosted years later by a mismatched vaccine, generating high titers of cross-reactive neutralizing antibodies. In our previous work, we observed that a primary vaccination series with two doses produced a response limited to the vaccine virus, but a later dose of a different vaccine formulation increased antibody levels against various viruses, including different clades. The observation that a second dose does not further increase antibody responses in previously vaccinated individuals aligns with our earlier findings^22^. In these individuals, the first dose likely already activated the B memory cell response, and a second dose shortly after may bind these antibodies rather than activate a new response.

The previously vaccinated participants did not have neutralizing antibodies at baseline (zero out of nine by MN, one out of nine by HI), even though binding antibodies were detected by FMIA. Since a single vaccination elicited high levels of neutralizing antibodies, it is reasonable to assume that the response targeted previously encountered epitopes (immunological recall), which are shared or cross-reactive among A(H5) antigens. Immune responses to influenza virus antigens are influenced by pre-existing immunity^25^, where primary encounters with an antigen shapes subsequent humoral and cellular immune responses, a phenomenon also known as imprinting^26^. In an epidemic situation, it may be beneficial to administer the two vaccine doses close together to achieve protection quickly. However, if the goal is to achieve the best possible cross-protection against viruses different from the vaccine strain, it might be more sensible to extend the interval between doses by several weeks or months. In recent years, particularly with COVID-19 vaccines, several studies have shown that the longer the interval between doses, the higher the antibody response achieved with the subsequent dose was^27,28^. Another long-term strategy could be to prime the at-risk population with a single dose of an H5 vaccine first, so that in a pandemic situation, only one booster vaccine would be needed to recall immunity.

In this study, we measured functional antibodies against the clade 2.3.4.4b A(H5N1) virus detected in cattle in the United States. This specific virus strain was isolated from the first human case during the early phase of the outbreak in March, 2024 (https://www.cdc.gov/han/2024/han00506.html)^29^. It is important to note that while we observed that the vaccine-induced antibodies effectively recognized this outbreak-related virus strain, it is possible that changes occurred in the virus over the course of 2024, including the HA protein, which may affect vaccine-induced protection. Mutations in the HA of the clade 2.3.4.4b viruses have been reported to occur in the HA head region, which includes the receptor-binding site and surrounding antigenic sites (https://cdn.who.int/media/docs/default-source/influenza/genetic-and-antigenic-characteristics-of-clade-2.3.4.4b-a(h5n1)-viruses-identified-in-dairy-cattle-in-the-united-states-of-america.pdf?sfvrsn=67aab002_3&download=true). The immunogenicity of the Zoonotic Influenza Vaccine Seqirus A(H5N8) was evaluated pre-clinically in a ferret model^11^. Cross-reactive responses were observed against different clade 2.3.4.4b strains, but no cross-reactivity was detected against A(H5) strains outside clade 2.3.4.4b. Additionally, no cross-reactivity was observed for a heterologous strain A/chicken/Ghana/AVL-76321VIR7050-39/2021 A(H5N1), despite it being within the same clade 2.3.4.4b as the vaccine. This strain possesses an HA A156T mutation predicting gained glycosylation in the antigenic site of HA, which may affect the recognition of antibodies raised by the zoonotic vaccine.

Constant evolution and diversification of the HA gene into different co-circulating phylogenetic groups is expected. Genomic monitoring of circulating and outbreak-associated viruses is crucial to evaluate vaccine recognition of contemporary viruses, potential cross-protective effects of the vaccine against clade 2.3.4.4b and for future vaccine composition recommendations in different geographic areas. Based on the observations in this study, we can conclude that the zoonotic influenza vaccine is expected to confer seroprotection against currently circulating H5 clade 2.3.4.4b viruses, with cross-protection despite ongoing antigenic drift. Given that a single dose induced high neutralizing antibody levels in subjects who had previously received vaccines based on H5 HA antigens from different clades, priming immunity in at-risk groups with the current vaccines may be beneficial for long-term heterologous immune memory. The low vaccination coverage among the target groups for whom the avian influenza vaccine is recommended underscores the necessity of targeted communication in the design of future vaccination campaigns. Even if the vaccine provides strong immune responses that matches with the circulating viruses, the ultimate determinant of its impact is the vaccination coverage within at-risk groups.

## Methods

### Study population and sampling

The “Avian and seasonal influenza vaccine induced immune responses” is a clinical study conducted in Finland by the Finnish Institute for Health and Welfare in collaboration with the Finnish Food Authority, HUS Diagnostic Center and University of Turku within the well-being services counties of Helsinki, Uusimaa, Kymenlaakso, Southern Carelia, Southern, Central and Northern Ostrobothnia and Kainuu (EU CT number 2023-509178-44-00) (https://euclinicaltrials.eu/search-for-clinical-trials/?lang=en&EUCT=2023-509178-44-00).

We invited individuals to whom the zoonotic influenza vaccine was recommended to participate in the study. Vaccination with the MF59-adjuvanted A(H5N8) influenza vaccine (clade 2.3.4.4b A/Astrakhan/3212/2020, Seqirus)^11^ was recommended to those at risk through direct or indirect exposure to infected animals including fur and poultry farm workers, veterinarians, bird ringers, and laboratory personnel handling the avian influenza virus or samples that may contain the virus. The national vaccination campaign started in Finland in June 2024. Vaccines were offered in accordance with the national recommendations given by the Finnish Institute for Health and Welfare (https://thl.fi/en/-/avian-influenza-vaccinations-begin-vaccine-to-be-offered-to-persons-at-increased-risk-of-infection) as a two-dose regimen with a minimum dose interval of three weeks. The vaccines were administered through routine healthcare services^9^.

The inclusion criteria for the study were (1) age of 18-65 years, (2) belonging to the target group of the avian influenza vaccine, (3) intention to accept at least one dose of the avian influenza vaccine, (4) a native speaker of Finnish, Swedish or English, (5) home address in Finland, (6) ability to give samples three weeks after each dose, (7) preferably also to participate in the follow-up samplings and (8) a written informed consent. The exclusion criteria were any medical contraindications to influenza vaccination and a history of anaphylactic reaction to any of the constituents or trace residues of the vaccine.

We invited all registered fur and poultry farmers in the wellbeing services counties of Southern, Central and Northern Ostrobothnia and Kainuu by mail. Farmers were asked to forward invitation letters to their employees. We approached public sector veterinarians, members of the national organization for bird conservation and birdwatching (BirdLife Finland) and laboratory workers at the Finnish Food Authority, Finnish Institute for Health and Welfare, Helsinki University Hospital and Diagnostic Center, Turku University Hospital and University of Turku, by sending an information letter of the study, and subsequently an invitation letter to those expressed their interest to participate in the study. Participants were asked to donate a blood sample at their local laboratory centre of the well-being services county at three study visits: baseline (maximum of 14 days before the first vaccine dose) and 18-24 days after the first and second vaccine dose. We included two cohorts in the study; (1) participants who belong to the target groups for which the avian influenza vaccine is recommended (targeted sample size 300) with no previous influenza (A)H5 vaccination history and (2) participants of cohort 1 who have previously received H5 influenza vaccines in 2009, 2011-2012 and /or 2018 (**Figure 1B**, **Table 1 and 2**).

The targeted sample size of 300 for the study cohort 1 was determined using the sample size formula: n= (Z2 x p(1-p))/E2. The calculation was based on a desired 95 % confidence level (Z), an assumed seroprotection rate of 75 % (p) and a 5 % margin of error (E). The result indicated a minimum sample size of 288 participants required to accurately estimate the proportion of subjects achieving seroprotection. With this sample size, the lower limit of the 95 % confidence interval is ≥70 %. The number of participants recruited to the study in 2024 remained significantly lower, which will introduce uncertainty into the seroprotection assessment in this study.

We retrieved contact information of fur and poultry farmers from the Central Database for Animal Keepers and Establishments maintained by the Finnish Food Authority. Information on avian influenza vaccinations given during the study was retrieved from the Register of Primary Health Care Visits. Participants were additionally asked to submit information on previous avian influenza vaccinations, which had in Finland been previously recommended for a limited target group of laboratory workers and veterinarians. The vaccines used in 2009, 2011-2012 and 2018 were the pre-pandemic A(H5N1), inactivated, AS03-adjuvanted A/Indonesia/5/2005 (clade 2.1.3.2)-like split virion vaccine, 3.75 µg HA (GlaxoSmithKline); A(H5N1), inactivated, adjuvant-free A/Vietnam/1203/2004 (clade 1)-like whole virus vaccine, 7.5 µg HA (Baxter) and A(H5N1), inactivated, MF59-adjuvanted A/turkey/Turkey/1/2005 (clade 2.2.1)-like strain (NIBRG-23) vaccine, 7.5 µg HA (Novartis), respectively.

Blood samples were collected from the participants at the three study visits at baseline and after each vaccine dose (**Figure 1B**). Serum was separated by centrifugation and stored at −20 °C until analyzed. All participants gave written informed consent before the collection of the first study sample.

### Cells

#### Cells for MN assay

Madin-Darby canine kidney (MDCK) cells (ATCC-CCL-34, lot 1805449) were maintained in Eagle’s Minimal Essential Medium with L-glutamine (L-Glu) and Earle’s balanced salt solution (EMEM, Gibco 6110087), containing 10 % fetal bovine serum (FBS, Sigma-Aldrich), 1 x non-essential amino acids (NEAA, Sigma-Aldrich), 1.1 g/l sodium hydrogen carbonate (CHNaCO_3_, Merck), 100 IU/ml penicillin (Pen, Sigma-Aldrich) and 100 mg/ml streptomycin (Strep, Sigma-Aldrich). Cells were tested mycoplasma negative, maintained at 37 °C, 5 % CO_2_ and passaged twice per week.

#### Cells for HI assay

MDCK cells (ATCC-CRL-2935) were maintained in EMEM (Capricorn Scientific) with Earle’s balanced salt solution, containing 10 % FBS, 1 x NEAA (Capricorn Scientific), 1.5 mg/ml sodium bicarbonate (NaHCO_3,_ Gibco), 10 mM 4-(2-hydroxyethyl)piperazine-1-ethane-sulfonic acid (HEPES, Capricorn Scientific), 100 IU/ml Pen (Capricorn Scientific), 100 mg/ml Strep (Capricorn Scientific) and 2 mM L-Glu (Capricorn Scientific). Human epithelial 293T cells (ATCC-CRL-3216) were maintained in Dulbecco Modified Eagle’s Medium, high glucose 4.5 g/l (DMEM, Capricorn Scientific) comprising 10 % FBS, 1 x NEAA, 1 mM sodium pyruvate (Gibco) supplemented with 2 mM L-Glu, 100 IU/ml Pen and 100 mg/ml Strep. Cells were tested mycoplasma negative, maintained at 37 °C, 5 % CO_2_ and passaged twice per week (MDCK cells when confluent and 293T cells when sub-confluent). For 293T cells, 500 mg/ml geneticin (Gibco) was added to the medium during basal cell culture.

### Viruses and antigens

Avian influenza virus strains used in MN and HI assays, and HA antigen used for FMIA are listed with background information in **Table 4**.

### Virus propagation for microneutralization assay

The A(H5N8) A/Astrakhan/3212/2020 candidate vaccine virus (CVV) with a modified protease cleavage site consistent with a low pathogenic phenotype (IDCDC-RG71A) was received by the Crick Worldwide Influenza Centre, London. The A(H5N1) A/blue fox/UH/004/2023 virus was isolated from a blue fox nasal sample from outbreak in fur animals in Finland in 2023^30^. The nasal sample was kindly provided by Associate Professor Tarja Sironen, University of Helsinki.

Virus strains used in MN test were further propagated in MDCK cells and harvested at the time of cytopathic effect between 50 and 75 %. A tissue culture infectious dose 50 % (TCID_50_) was determined and calculated by the Reed-Muench method for each virus stock separately as previously described^31^ using the same modified protocol as in the MN assay represented below.

### Generation of plasmids and recombinant viruses for hemagglutination inhibition assay

#### Plasmids

The A(H5N1) A/Texas/37/2024 virus was isolated from a dairy farm worker in the USA during the cattle outbreak in 2024 and plasmids were kindly provided by Dr. Daniel R. Perez, University of Georgia, the United States^32^. The HA segment of A(H5N8) A/Astrakhan/3212/2020 was synthesised by Proteogenix. The HAs were cloned into a reverse genetics plasmid (modified version of pHW2000) as described previously ^33^ by seamless cloning using the GeneArt^TM^ Seamless Cloning kit (Thermo Fisher Scientific).

#### Recombinant virus production and sequencing

Recombinant viruses were produced using the eight-plasmid rescue system as described previously^33^. For the HI assay, recombinant viruses carrying seven gene segments of A/Puerto Rico/8/1934 (PR/8) high yield (HY)^34^ and the A(H5) HA segment of interest, without the multibasic cleavage site (MBCS), were generated under biosafety level 2 (BSL2) conditions. Following virus rescue, virus production was evaluated by an hemagglutination (HA) assay with 1 % turkey red blood cells (tRBCs) in phosphate buffered saline (PBS). Virus stocks were propagated in MDCK cells twice and HA gene sequences were checked by Sanger sequencing using the 3500xL Genetic Analyzer (Applied Biosystems). Accession numbers can be found in **Table 4**.

### Microneutralization assay

An enzyme-linked immunosorbent assay (ELISA)-based MN assay was performed as previously described^21,22,31^, yet further optimized conjugate and substrate steps were used in this study. Duplicate (technical replicate) heat-inactivated (56 °C, 30 minutes) serum samples were two-fold serially diluted starting at 1:10 dilution in MN medium comprising of OptiPro^TM^ SFM medium (Gibco), supplemented with 0.2 % of bovine serum albumin (BSA), 1 x NEAA, Pen and Strep in a total volume of 50 μl. Equal volume of pre-titrated virus was added to obtain 100 × TCID_50_ per well, following incubation for one hour at 37 °C, 5 % CO_2_. MDCK cells were detached, counted, and added in a total volume of 100 μl (2.5 x 10^4^ cells/well), and the 96-well flat base tissue culture plates (Sarstedt) were incubated at 37 °C, 5 % CO_2_ for 18-20 hours. Wells were washed once with PBS and fixed with ice cold 80 % acetone for 10 minutes.

The presence of influenza A virus in infected cells was detected by ELISA. Fixed plates were washed twice with washing buffer consisting of PBS containing 0.05 % Tween® 20 (Sigma-Aldrich). A horseradish peroxidase-labeled (HRP Conjugation Kit - Lightning-Link®, Abcam) influenza A nucleoprotein-specific antibody (A7307, Medix Biochemica) was diluted 1:10,000 in PBS containing 5 % milk and incubated (80 μl/well) at room temperature for one hour. After washing six times with the washing buffer, 100 μl of substrate (1-Step™ TMB ELISA Substrate Solutions, Thermo Scientific™) was added into each well and incubated at room temperature for 20 minutes in the dark. The reaction was stopped with 100 μl of 2 N sulfuric acid. Absorbances were measured within 30 minutes at 450 nm and 620 nm.

The neutralizing endpoint was determined as previously described^31^. Results were expressed as titers corresponding to the reciprocal of the serum dilution that inhibited 50 % of influenza infection. MN titer ≥10 was considered positive, and negative when <10. If the titer was <10, a titer of 5 was assigned as the result.

### Hemagglutinin inhibition assay

Recombinant avian influenza viruses in the PR/8 HY background were tested using horse red blood cells (hRBCs) obtained from Cerba Research, Rotterdam, the Netherlands. hRBCs were used instead of the more commonly utilized tRBCs, due to their nearly exclusive expression of α2,3-sialic acid (SA) receptors on their surface, which are preferentially bound by avian influenza viruses^16^.

After collection, horse blood in citrate buffer was stored at 4 °C for up to one month. Prior to use, hRBCs were washed three times by centrifugation at 1800 rpm (acceleration: 9 and deceleration: 2) with PBS for 10 minutes at room temperature. Final concentrations of 2 % and 10 % hRBCs in PBS were made.

Serum samples were absorbed with an equal volume of 10 % hRBCs at 4 °C for one hour with mixing every 20 minutes, to prevent nonspecific agglutination. Subsequently, nonspecific inhibition was avoided by incubating sera with in-house manufactured *Vibrio cholerae* filtrate comprising receptor destroying enzyme (RDE) at a 1:6 ratio (volume/volume) overnight at 37 °C following RDE inactivation at 56 °C for one hour.

Post RDE inactivation, two-fold serial dilutions of sera in 0.5 % BSA (Sigma-Aldrich) in PBS (0.5 % BSA-PBS) were prepared in 96-well V-bottom microtiter plates (Greiner) starting at a 1:20 dilution in a total volume of 50 μl. Viruses were adjusted to 4 hemagglutinating units (HAU) per 25 μl in PBS and added to each well. Plates were mixed and incubated at 37 °C for 30 minutes. Following this, 25 μl of 2 % hRBCs was transferred to each well, plates were tapped individually, and HI titers were determined after a 1.5-hour incubation at 4 °C. In case of agglutination in the serum control well(s), the HI assay with the corresponding sera was repeated. Six serum samples were absorbed twice instead of once to remove nonspecific agglutination. The HI titers were defined as the inverse of the last serum dilution in which hRBC agglutination was partially or completely inhibited. The detection limit entailed an HI titer of 10, which was assigned to those serum samples that revealed partial agglutination in the first well. If the titer was <10, a titer of 5 was assigned as the result. Data are presented as a single experiment.

### Binding antibodies measured with fluorescent bead-based multiplex immunoassay

The binding of serum IgG to A(H5) was measured with FMIA adapted from an assay used in detection of SARS-CoV-2 antibodies^35^. A 100 µg/ml of A(H5N1) A/Michigan/90/2024 HA (REC32116, Native Antigen Company) was conjugated onto MagPlex-C superparamagnetic carboxylated beads (Luminex). Subsequently, 25 µl of beads diluted in PBS (pH 7.2) were added to black 96-well flat base plates (Costar 3915, Corning) with 25 µl of serum diluted in PBS (pH 7.2 with 1 % BSA, 0.8 % polyvinylpyrrolidone, 0.5 % poly(vinyl alcohol) and 0.1 % Tween-20). The plates were incubated for one hour. This and all subsequent incubations were performed at room temperature in the dark with shaking at 600 rpm. After washing with a magnetic plate washer (405TSRS, BioTek), 50 µl of 1:100 diluted IgG detection antibody (R-phycoerythrin-conjugated AffiniPure goat anti-human IgG Fcγ fragment-specific detection antibody, Jackson ImmunoResearch) was added and plates were incubated for 30 minutes. Following washing, 80 µl of PBS (pH 7.2) was added and plates were incubated for 5 minutes. Fluorescence was measured with MAGPIX® System (Luminex). Median fluorescent intensity was converted into FMIA U/ml by interpolation from 5-parameter logistic curves (xPONENT v.4.2, Luminex) created from serially diluted (1:400-1:1638400) in-house reference pooled from this study’s sera. All plates were run with duplicates of in-house reference, blank and two control samples. All samples were analysed diluted 1:400 and 1:1600 in duplicate, and result was calculated as an average of four wells. Samples with fluorescence exceeding reference serum’s linear area were reanalysed more diluted.

### Ethical statement

The study was conducted according to the guidelines of the Declaration of Helsinki and was submitted for evaluation through the EU Clinical Trial Information System. The study has been approved by the National Committee on Medical Research Ethics (decision number TUKIJA/7/2024) and received authorization from the Finnish Medicines Agency Fimea (EU CT number 2023-509178-44-00).

### Statistical methods

All statistical analyses were performed with GraphPad Prism version 10.2.3 and R version 4.2.1. *P* values <0.05 were considered statistically significant. For intra- and inter-group comparison data was categorized into two groups according to vaccination history. Only data from participants who provided samples at the three different time points and received vaccinations in the correct order relative to sampling were included in the analysis of vaccine responses. Geometric means and 95% confidence intervals were calculated for the neutralizing antibody titers and IgG antibody concentrations. Fold changes were calculated from geometric means of each group.

Since MN and HI data were not normally distributed, the data was log2-transformed and non-parametric tests were used. Mann–Whitney U test was used to compare differences between different groups, while the Wilcoxon matched-pairs signed-rank test was used within the group comparisons across different time points. As IgG data was normally distributed, comparisons between groups were performed with T-test.

To assess the correlation of titers against A/Astrakhan/3212/2020 measured with MN and HI, data from all 142 samples that had results measured with both tests, including serum samples from participants who did not provide all 3 samples, were included. An HI titer of 40 is typically accepted to correspond to a 50 % or more reduction in the risk of contracting an influenza infection or influenza disease^36^ and defined by both the U.S. Food and Drug Administration and the European Medicines Agency Committee for Medicinal Products for Human Use as the primary correlate of protection^37^. To determine the MN titer corresponding to an HI titer of 40 against A/Astrakhan/3212/2020, the data was log2-transformed, and Spearman’s correlation was performed (r = 0.8836, *p* <0.0001). The Spearman correlation coefficient indicated a positive correlation between MN and HI titers (Figure 2F). To further explore this relationship, regression analysis was conducted. A simple linear regression model was applied to assess the equivalence between MN and HI titers (R^2^ = 0.7614, *p* <0.0001) yielding the equation Y = 0.9098x-0.4967. Based on this model, an HI titer of 40 corresponds to an MN titer of 20. The percent seroprotection rate (SPR %) for each group was calculated as number of seropositive samples (MN titers ≥20 or HI titers ≥40) divided by the number of samples x 100 in the group. Confidence intervals for SPRs were calculated with normal approximation to the binomial calculation.

## Data availability

Data subject to third party restrictions. The data that support the findings of this study are available from Finnish Social and Health Data Permit Authority Findata. Restrictions apply to the availability of these data, which were used under license for this study under informed consent form. Anonymized data are available after permission by Findata.

## Acknowledgements

We thank Saara Suopanki, Lotta Hagberg, Maria Heikkilä, Raisa Hanninen and Marja Suorsa for handling of clinical documents and samples, Maria Heikkilä, Saara Suopanki and Mikko Määttä for performing laboratory analyses, Tarja Sironen, Veera Avelin, Eda Altan and Pamela Österlund for A/blue fox/UH/004/2023 virus isolation, Marja-Liisa Ollonen, Kirsi Mäkisalo and Mervi Eskelinen for cell culture, Mikko Aura, Esa Ruokokoski, Juha Oksanen and Teemu Rauhala for data management, Joni Niemi for sample management and Carita Savolainen-Kopra, Anna Katz, Otto Helve from THL and Marion Koopmans from Erasmus MC for providing the necessary facilities and resources that enabled us to conduct the research effectively. We thank Arto A. Palmu, Elina Isosaari and Heta Nieminen from FVR – Finnish Vaccine Research for guidance on the conducting of the clinical study, study coordination and regulatory processes. We gratefully acknowledge the authors and their respective laboratories, who analyzed and submitted the sequences to GISAID’s EpiFlu^TM^ Database. In addition, we would like to thank all the laboratory centers and volunteers who took part in this study.

## Author contributions

Merit Melin, Hanna Nohynek, Ritva Syrjänen, Nina Ekström, and Ilkka Julkunen designed the study. Oona Liedes, Anu Haveri, Anna Solastie, Willemijn F. Rijnink and Theo M. Bestebroer conducted the immunological research. Oona Liedes, Anu Haveri, Anna Solastie, Willemijn Rijnink, Merit Melin, Mathilde Richards, and Rory de Vries contributed to the data analysis. Nina Ekström and Ritva Syrjänen managed the coordination and regulatory process of the clinical study. Nina Ekström, Terhi Laaksonen, Riikka Holopainen, Laura Kakkola, and Maija Lappalainen contributed to the recruitment. Oona Liedes, Anna Solastie, Anu Haveri and Saimi Vara were involved with handling of the clinical documents and samples. Merit Melin, Oona Liedes, Nina Ekström, Anna Solastie, Anu Haveri, Saimi Vara, Willemijn Rijnink, Rory de Vries, Erika Lindh and Hanna Nohynek wrote the manuscript. Terhi Laaksonen, Riikka Holopainen, Maija Lappalainen, Ilkka Julkunen, Laura Kakkola, and Ritva Syrjänen, Niina Ikonen and Mathilde Richard assisted with the editing of the text.

